# Cardiometabolic risks of SARS-CoV-2 hospitalization using Mendelian Randomization

**DOI:** 10.1101/2020.12.30.20248889

**Authors:** Noah Lorincz-Comi, Xiaofeng Zhu

**Affiliations:** Department of Population and Quantitative Health Sciences, Case Western Reserve University, Cleveland, OH, USA

## Abstract

**Intro:** Many cardiometabolic conditions have demonstrated associative evidence with COVID-19 hospitalization risk. However, the observational designs of the studies in which these associations are observed preclude causal inferences of hospitalization risk. Mendelian Randomization (MR) is an alternative risk estimation method more robust to these limitations that allows for causal inferences.

**Methods & materials:** We applied four MR methods (MRMix, IMRP, IVW, MREgger) to publicly available GWAS summary statistics from European (COVID-19 GWAS n=2,956) and multi-ethnic populations (COVID-19 GWAS n=10,808) to better understand extant causal associations between Type II Diabetes (GWAS n=659,316), BMI (n=681,275), diastolic and systolic blood pressure, and pulse pressure (n=757,601 for each) and COVID-19 hospitalization risk across populations.

**Results:** Although no significant causal effect evidence was observed, our data suggested a trend of increasing hospitalization risk for Type II diabetes (IMRP OR, 95% CI: 1.67, 0.96-2.92) and pulse pressure (OR, 95% CI: 1.27, 0.97-1.66) in the multi-ethnic sample.

**Conclusions:** Type II diabetes and Pulse pressure demonstrates a potential causal association with COVID-19 hospitalization risk, the proper treatment of which may work to reduce the risk of a severe COVID-19 illness requiring hospitalization. However, GWAS of COVID-19 with large sample size is warranted to confirm the causality.

## Introduction

SARS-CoV2-2, a novel coronavirus (COVID-19), has infected millions of individuals globally since it first emerged in December 2019. There is tremendous effort to learn more about why some individuals progress from a relatively stable illness to a more serious one requiring hospitalization. Observational studies, while providing some evidence for the conferral of hospitalisation risk by a number of risk factors, have sometimes produced conflicting results^[1]^. Equally, it would be inappropriate to infer from these studies alone causal associations between a given exposure and COVID-19 hospitalization. The explanations for this vary but may include unresolved bias, confounding, and reverse causation^[2,3]^. One alternative to observational studies is the randomised controlled trial (RCT) that can effectively eliminate confounding and reverse causation effects via the randomisation process. However, an RCT to identify causal risks for COVID-19 hospitalization would be expensive and unfeasible because it would be impossible to randomize patients based on a hypothesized risk factor in practice. Mendelian randomisation (MR) is an alternative to the RCT that retains the randomisation component but is both feasible and inexpensive^[4]^. MR capitalizes on the randomisation of an individual’s genetic information during meiosis. This means that the effects of each genetic variant on a given outcome should be less subject to the effects of residual confounding, such as reverse causation since genes are at conception fixed. This makes MR extremely powerful, which could explain its recent popularity in biomedical research.

Observational epidemiological studies have identified Type II Diabetes, hypertension, and body mass index (BMI) as potential risks for COVID-19 hospitalization (vs non-hospitalization). Pooled estimated odds ratios are 2.75 (95% CI: 2.09-3.62) for Type II diabetes^[5]^, 2.30 (1.76-3.00) for hypertension^[6]^ (binary), and 2.67 (1.52-3.82) for BMI^[7]^. Not only did all these pooled estimates display heterogeneity in the meta-analysis from which they came, the observational designs of the included studies preclude making any causal inferences. We attempt here to further investigate some of these associations using MR, namely the associations between Type II Diabetes, body mass, diastolic and systolic blood pressure, and pulse pressure. We test for the causal relationships between each of these health conditions and COVID-19 severity, as indicated by hospitalization risk, using publicly available summary statistics from genome-wide association studies (GWAS). These exposures were selected for their hypothesized associations with COVID-19 hospitalization.

## Methods

### Data

COVID-19 summary statistics were retrieved from the COVID-19 Host Genetics Initiative^[8]^ (covid19hg.rg/results) for 928 European cases and 2,028 controls and 2,430 cases and 8,378 controls of European, African, Hispanic, and Middle Eastern ethnicity. Cases were those hospitalised COVID-19 patients and controls were non-hospitalised COVID-19 patients. GWAS estimates, from three research groups^[9-11]^ (UK Biobank, deCODE Genetics, FinnGen), of European individuals were meta-analyzed to yield the complete set of summary statistics for Europeans. GWAS estimates from 10 additional research groups^[12-19]^ (BoSCO, SPGRX, GNH, PMBB, QGP, MVP, Ancestry, BQC19; full descriptions of these groups are found at the respective links in the References section) were then analysed with the original three European cohorts to form the expanded set of estimates from more ethnically diverse individuals.

GWAS summary statistics from three European descent studies were meta-analysed using inverse variance-weighting approach to yield a single set of summary statistics for 12,029,423 and 9,503,351 SNPs for European and variable ethnicity populations, respectively. Only alleles with frequency >0.1% were included in the meta-analysis. Summary statistics for all five exposures, namely diastolic and systolic blood pressure (DBP, SBP), pulse pressure (PP), body mass index (BMI), and Type II diabetes (T2D), were available from public GWAS repositories. We used PLINK v1.90b6.11^[20]^ to select independent GWAS significant SNPs from the available millions of SNPs for each exposure. These SNPs had in their GWAS a p-value less than 5e-8 and a linkage disequilibrium coefficient less than 0.1. For BMI (n=681,275)^[21]^, DBP, SBP, PP (n=757,601 for each)^[22]^, and Type II Diabetes (n=659,316)^[23]^ 1,494, 1,202, 1,134, 926, and 174 SNPs meeting these criteria were identified, respectively. The GWAS data repositories for each exposure are listed in the respective referenced research.

For each exposure, we merged the COVID-19 summary statistics to each exposure data set by matching genetic variant and effect alleles between the exposure and COVID-19 set of summary statistics. No SNPs in the original COVID-19 GWAS had between-study heterogenous effect estimates significant at a Bonferroni-adjusted p-value threshold. In the set of summary statistics from only European cohorts, 129, 1,186, 947, 884, 723 selected SNPs (using selection procedures described above) were included in the analyses for Type II Diabetes, BMI, DBP, SBP, and PP, respectively; in cohorts of individuals of variable ethnicity, 129, 1,188, 953, 887, and 726 SNPs were selected for Type II Diabetes, BMI, DBP, SBP, and PP, respectively. Selected SNP counts different between samples because the COVID-19 GWAS SNP counts differed between samples from the beginning.

### Mendelian Randomization Analysis

To estimate the causal effects of each exposure on the probability of COVID-19 hospitalization status, we used Mendelian Randomization (MR). This method intends to eliminate potential confounding effects by capitalizing on the random assignment of genetic information at conception. The causal effect of each variant *i* is generally estimated as 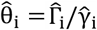 where 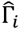 and 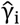 are the standardized effect estimates of SNP *i* on the outcome and exposure, respectively. Standardized effect estimates are calculated as

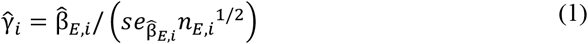

for SNP *i* of the exposure (*E*) and

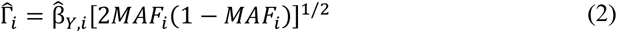

for the outcome *Y* where *MAF*_*i*_ is the minor allele frequency. The standard errors of the effect sizes are also adjusted from the original standard errors in each GWAS as:

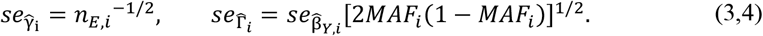

Many methods for estimating the causal effect exist, of which we chose four that are: MRMix^[24]^, IVW^[25]^, MR-Egger^[26]^ and IMRP^[27]^. Comparing the results of four similar but distinct analytical procedures provided a more complete picture of any existing causal relationships between each exposure and COVID-19 hospitalization. All analyses are more fully described elsewhere^[28]^ and were done using the *MRMix* and *IMRP* packages in R^[29]^.

## Results

Table 1 presents summary statistics for the effect sizes of each exposure and for the correlations between GWAS effect estimate t-statistics for each exposure and COVID-19. These summary statistics indicate that, for each exposure in European only or multiple ethnic sample, the mean exposure effect sizes of only the selected SNPs are all <0.001. In the European-only sample, the correlations of effect sizes for the respective exposure and COVID-19 hospitalization [i.e.,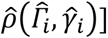 are all negative. In the multiple ethnicity sample, the correlations are more robust. These associations are reflected in Figure 1.

**Table 1:** Summary statistics for exposure effect sizes

|  | Effect size |  |  |  |  |  |  |  |  |  |
| --- | --- | --- | --- | --- | --- | --- | --- | --- | --- | --- |
|  | Europeans<sub>a</sub> |  |  |  |  | Multiple Ethnicity<sub>a</sub> |  |  |  |  |
| Exposure | Mean | SD | Min. | Max. | $\hat{\rho}$ (exposure, COVID-19)<sub>b</sub> (SE) | Mean | SD | Min. | Max. | $\hat{\rho}$ (exposure, COVID-19)<sub>b</sub> (SE) |
| Type II Diabetes | <0.001 | 0.010 | -0.021 | 0.020 | -0.033 (0.089) | <0.001 | 0.010 | -0.021 | 0.020 | 0.153 (0.084) |
| BMI | <0.001 | 0.009 | -0.022 | 0.043 | -0.024 (0.029) | <0.001 | 0.009 | -0.022 | 0.043 | 0.013 (0.029) |
| DBP | <0.001 | 0.009 | -0.025 | 0.023 | -0.035 (0.033) | <0.001 | 0.009 | -0.025 | 0.023 | 0.026 (0.032) |
| SBP | <0.001 | 0.009 | -0.026 | 0.023 | -0.027 (0.034) | <0.001 | 0.009 | -0.026 | 0.023 | 0.036 (0.033) |
| PP | <0.001 | 0.009 | -0.022 | 0.027 | -0.021 (0.037) | <0.001 | 0.009 | -0.022 | 0.027 | 0.089 (0.037) |

**a:** Estimates from the European set are from 2,956 European individuals; estimates from the variable ethnicity set are from 10,808 individuals of either of European, African, Hispanic, or Middle Eastern descent. **b:** the correlation between the effect sizes for each SNP's estimated effect on the probability of the respective exposure or COVID-19 hospitalization after all mentioned exclusions (SNP independence, $p < 5e-8$ , Pearson's $r < 0.1$ with index SNP) were applied.

**Figure 1:**
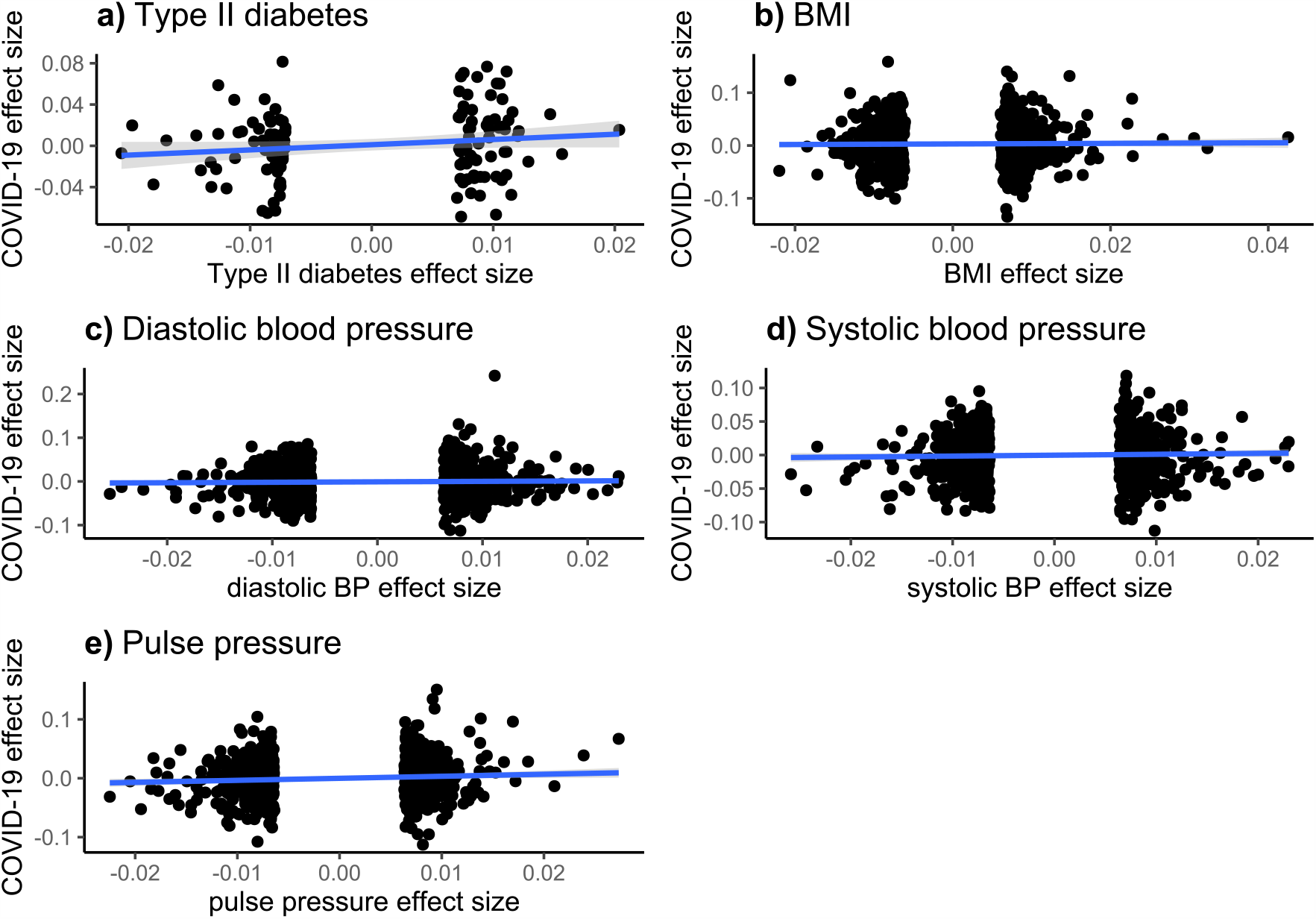
Exposure, COVID-19 Effect Size Associations – Mixed Ethnicity Sample. These figures display the effect size associations between each exposure (Type II Diabetes, BMI, diastolic and systolic blood pressure, and pulse pressure) and COVID-19 hospitalization risk. Overlaid on the scatterplots are univariate linear regression fitted values and their associated 95% confidence intervals.

Using the summary statistics from only European individuals, we did not observe significant causal effects of the exposures to COVID-19 hospitalization after adjusting for multiple tests, although a nominal evidence for BMI was observed in the IMRP analysis (OR=0.66, 95% CI: 0.46-0.96). No uniformly significant causal effect evidence was observed in the multiple ethnicity sample although all the exposures demonstrated increasing risk of COVID-19 hospitalization (Table 2). In the sample of individuals of multiple ethnicities, Type II Diabetes was positively associated with COVID-19 hospitalization using IMRP (1.672, 0.956-2.92). Positive associations are also detected by MRMix, IVW, and MREgger models, though with attenuated effect estimates and wider confidence intervals. Using IMRP, six of the 129 variants in the Type II Diabetes analyses displayed evidence of pleiotropic effects (P<0.05) which are shown in Figure 2, although none remain after Bonferroni correction. The intercept term in the MREgger model was non-significant (p=0.784), indicated an absence of pleiotropic effects or low statistical power for detecting them.

**Table 2:**
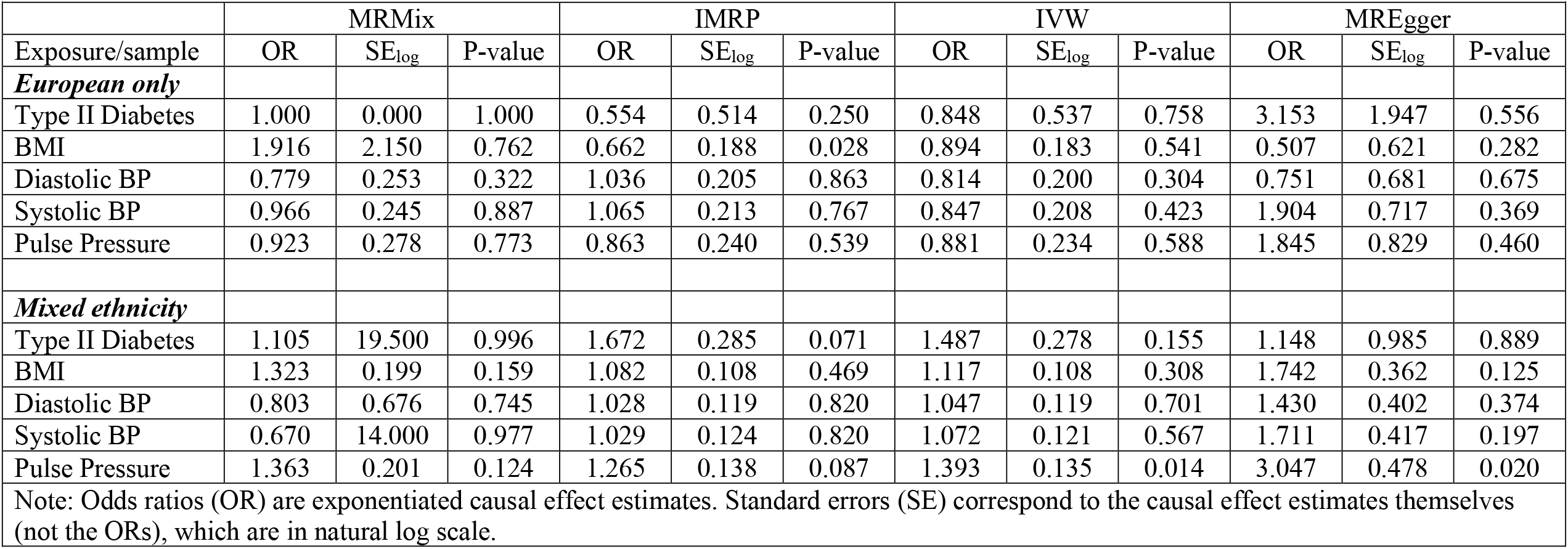
Causal effect estimates

**Figure 2:**
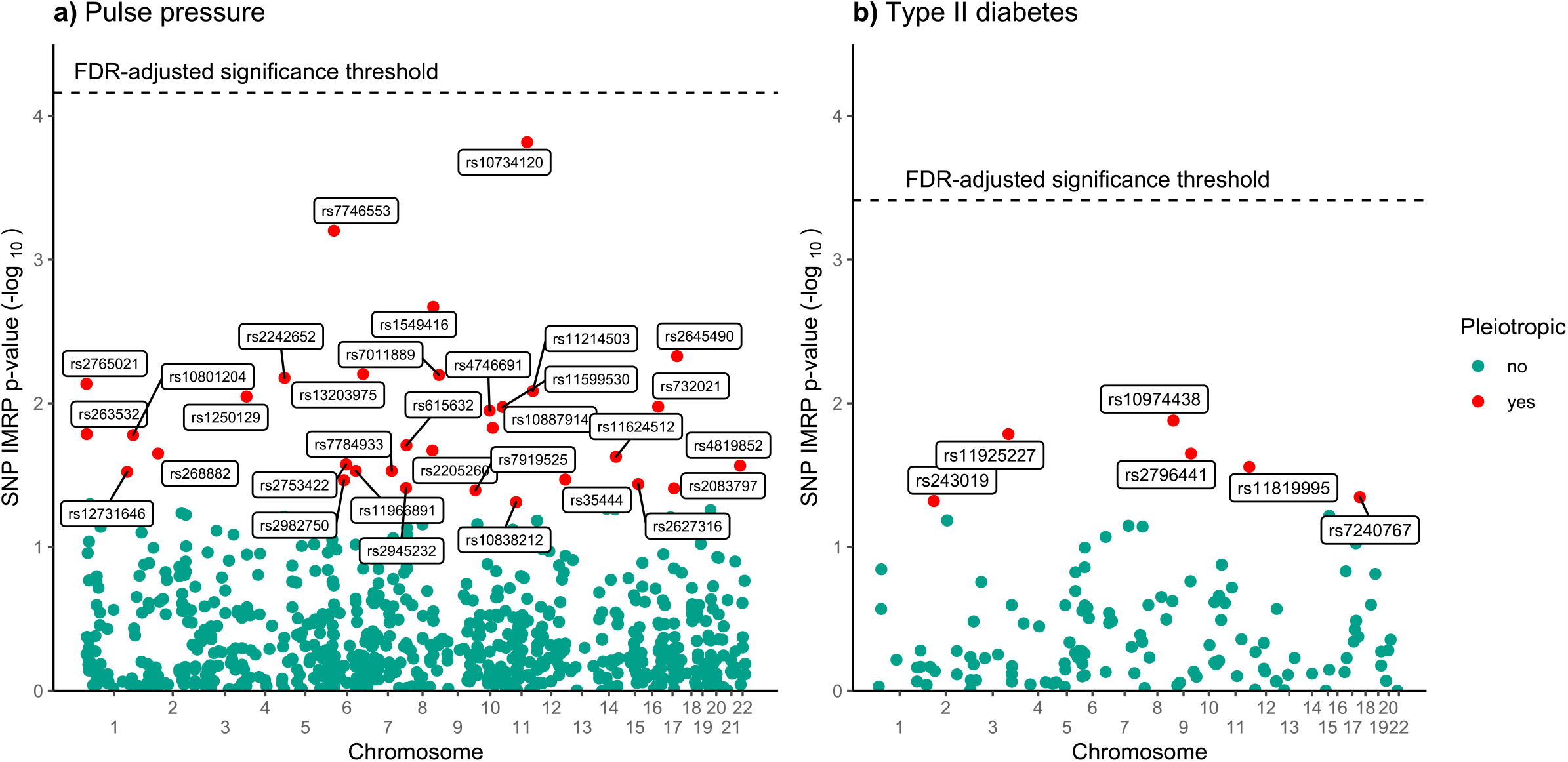
IMRP Pleitropy Evidence – Mixed Ethnicity Sample. Displayed are those SNPs demonstrating evidence of pleiotropic effects for pulse pressure and Type II Diabetes in the multi-ethnicity sample as estimated by IMRP. The false discovery rate-adjusted threshold is determined by a Bonferroni-adjusted Type I Error rate of p=0.05.

A positive causal association was detected for pulse pressure in the multiple ethnicity sample. The IMRP model produced an estimated odds ratio of 1.27 (95% CI: 0.97-1.66). Positive associations are also detected by IVW (1.39, 1.07-1.81) and MREgger (3.05, 1.20-7.77), and marginally so for MRMix (1.36, 0.92-2.02). Thirty-two SNPs displayed evidence of pleiotropic effects (P<0.05) as indicated by IMRP (SNPs included in Figure 2, detailed in Supplementary Table S1), none of which remained after a Bonferroni adjustment (MREgger intercept p-value: 0.088). To better understand why for diastolic and systolic BP in the mixed ethnicity sample MRMix produced a causal effect estimate in the direction opposite of that produced by IMRP, IVW, and MREgger, we also plotted the causal effect estimate 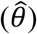 against its likelihood function (see Figure 3). For both variables a clear but flat peak is present, suggesting large standard error. A possible reason is the small sample size of COVID-19.

**Figure 3:**
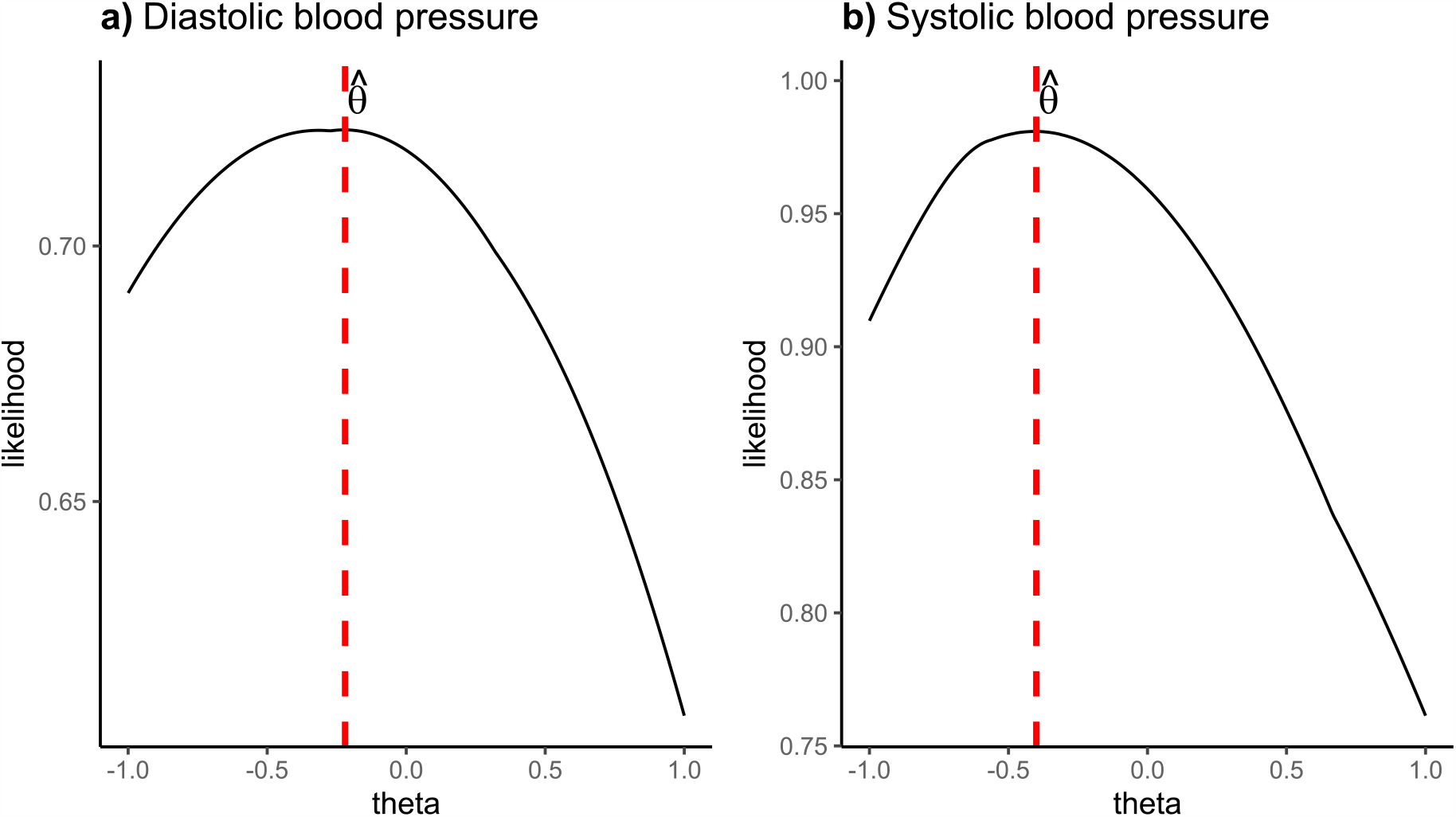
MRMix Estimation Performance – Mixed Ethnicity Sample. These two plots display the estimated causal effect (theta,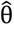) during maximum likelihood estimation for diastolic and systolic blood pressure by MRMix in the multi-ethnicity sample. A clear, sharp peak indicates stable performance in the estimation of theta. The dotted red lines indicate the most likely estimates of theta (θ) reported in the Results section, respectively for each exposure.

## Discussion

Using summary statistics from European-only samples, we failed to detect any compelling evidence supporting any causal associations between hospitalization from COVID-19 and Type II Diabetes, body mass index, diastolic and systolic blood pressure, and pulse pressure. Conversely, we provide evidence of existing positive causal associations between Type II Diabetes and pulse pressure and the probability of hospitalization from COVID-19. A positive causal association between pulse pressure and COVID-19 hospitalization is biologically plausible as pulse pressure is moderately correlated with lung function (r=-0.37)^[30]^, the failure of which is a primary reason for COVID-19 hospitalization. Similarly, although Type II diabetes is associated with a number of risks also hypothesized to be causally associated with COVID-19 severity (e.g., obesity, hypertension, cardiovascular disease), risk for hospitalization may also be conferred because of known inflammatory immune responses in patients with Type II Diabetes^[31,32]^.

Nonetheless, our study is limited by many factors, not least of which is the relatively small sample sizes from which the COVID-19 hospitalization summary statistics were produced (n=2,956 for only Europeans, 10,808 for variable ethnicities). While we cannot for certain say that this alone can explain some of the null findings, using COVID-19 summary statistics from even larger GWAS samples would have allowed for more accurate estimates of any extant causal associations between each exposure and hospitalization from COVID-19. As more GWAS using COVID-19 patients are completed, more accurate summary statistics produced from larger samples will become publicly available and thus future work will work to reduce the probability of making Type II errors.

Conversely, our study is strengthened by the very large sizes of the samples from which the exposure summary statistics were drawn. Each of these GWAS included >650,000 participants. Equally, our study is strengthened by the availability of GWAS summary statistics from different populations of individuals. This of course may allow us to generalize our findings to more people globally. Lastly, the comparison of results across multiple causal effect estimation methods should provide greater confidence in the findings for each exposure. We also benefit from a relatively new method (IMRP) which can detect SNPs displaying evidence of pleiotropic effects. Future work should build on this research to better understand COVID-19 disease burden within and across many populations so as to minimize national and global hospitalization risk.

## Data Availability

All data used in this study are publicly available from the author references cited in the Data subsection of the Methods section.

## Acknowledgements

This work was supported by grant HG011052 (to X.Z.) from the National Human Genome Research Institute (NHGRI). N. L-C. was supported by grants T32 HL007567 from the National Heart, Lung and Blood Institute (NHLBI).

## Disclosures

There are no conflicts of interest.

## Notes

### Competing Interest Statement

The authors have declared no competing interest.

### Author Declarations

The study was approved by the Institute Review Board (IRB) at Case Western Reserve University (IRB number: STUDY2018059).

