## Supplemental Table 1 for "Cardiometabolic risks of SARS-CoV-2 hospitalization using Mendelian Randomization"

**Supplementary Table S1: Pleiotropy by SNP for Type II Diabetes, Pulse Pressure**

| Exposure | SNP | P-value |
| --- | --- | --- |
| ***Type II Diabetes*** |  |  |
|  | rs10974438 | 0.013169 |
|  | rs11925227 | 0.016348 |
|  | rs2796441 | 0.022327 |
|  | rs11819995 | 0.027647 |
|  | rs7240767 | 0.044900 |
|  | rs243019 | 0.047797 |
| ***Pulse Pressure*** |  |  |
|  | rs10734120 | 0.000152 |
|  | rs7746553 | 0.000629 |
|  | rs1549416 | 0.002129 |
|  | rs2645490 | 0.004694 |
|  | rs13203975 | 0.006249 |
|  | rs7011889 | 0.006338 |
|  | rs2242652 | 0.006652 |
|  | rs2765021 | 0.007312 |
|  | rs11214503 | 0.008208 |
|  | rs1250129 | 0.008968 |
|  | rs732021 | 0.010537 |
|  | rs11599530 | 0.010602 |
|  | rs4746691 | 0.011229 |
|  | rs10887914 | 0.014797 |
|  | rs263532 | 0.016353 |
|  | rs10801204 | 0.016618 |
|  | rs615632 | 0.019559 |
|  | rs2205260 | 0.021249 |
|  | rs268882 | 0.022319 |
|  | rs11624512 | 0.023533 |
|  | rs2753422 | 0.026566 |
|  | rs4819852 | 0.027124 |
|  | rs11966891 | 0.029493 |
|  | rs7784933 | 0.029511 |
|  | rs12731646 | 0.029949 |
|  | rs35444 | 0.033905 |
|  | rs2982750 | 0.034263 |
|  | rs2627316 | 0.036398 |
|  | rs2945232 | 0.038756 |
|  | rs2083797 | 0.038998 |
|  | rs7919525 | 0.040251 |
|  | rs10838212 | 0.048706 |

These P-values are from fitted IMRP models using summary statistics from the sample of individuals of multiple ethnicities.
